# Regional disparities in Dementia-free Life Expectancy in Japan: an ecological study, using the Japanese long-term care insurance claims database

**DOI:** 10.1101/2022.12.27.22283979

**Authors:** Mikako Yoshikawa, Etsu Goto, Jung-ho Shin, Yuichi Imanaka

## Abstract

**Background:** The number of people with dementia increases in an aging society; therefore, promoting policies for dementia throughout the community is crucial to creating a dementia-friendly society. Understanding the status of older adults with dementia in each region of Japan will be a helpful indicator. We calculated Dementia-free Life Expectancy and aimed to examine regional disparities and their associated factors.

**Methods:** We calculated Dementia-free Life Expectancy and Life Expectancy with Dementia for each secondary medical area in Japan based on the Degree of Independence in Daily Living for the Demented Elderly, using data extracted from the Japanese long-term care insurance claims database. We then conducted a partial least squares regression analysis, the objective variables being Dementia-free Life Expectancy and Life Expectancy with Dementia for both sexes at age 65, and explanatory regional-level variables included demographic, socioeconomic, and healthcare resources variables.

**Results:** The mean estimated regional-level Dementia-free Life Expectancy at age 65 was 17.33 years (95% confidence interval [CI] 17.27–17.38) for males and 20.05 years (95% CI 19.99–20.11) for females. Three latent components identified by partial least squares regression analysis represented urbanicity, socioeconomic conditions, and health services-related factors of the secondary medical areas. The second component explained the most variation in Dementia-free Life Expectancy of the three, indicating that higher socioeconomic status was associated with longer Dementia-free Life Expectancy.

**Conclusions:** There were regional disparities in secondary medical area level Dementia-free Life Expectancy. Our results suggest that socioeconomic conditions are more related to Dementia-free Life Expectancy than urbanicity and health services-related factors.

## Introduction

Today, the world’s population is aging, especially in developed countries, and the number of people with dementia is increasing [1]. Japan is one of the most aged countries, and it has the highest prevalence of dementia (23.3 per 1,000 people) in the world [2], which burdens people with dementia, their communities, and the nation as a whole [3, 4]. With the growing importance of addressing the social problems of dementia, the Japanese government released the “Framework for Promoting Dementia Care” in 2019 [5]. It aims to delay the onset of dementia and create a society where people can live their daily lives with hope, regardless of whether they have dementia. This framework also mentions the need to collect and analyze evidence of effective measures to promote at the community level. Many studies have reported various healthy life expectancies to assess a population’s health status [6, 7]. For example, dementia-free life expectancy has been calculated in several countries via different methods, revealing that a healthy lifestyle is associated with a larger proportion of remaining years without dementia [8-11]. Those studies calculated dementia-free life expectancy by predicting the country’s prevalence from extracted data, but few have clarified differences among smaller units or regions within a country [12].

Under Japan’s Long-term care insurance (LTCI) system, people aged 65 years or older and persons aged 40 to 65 years with the specified conditions receive a “long-term care need certification” to utilize LTCI services such as home-visit care and daycare services. The level of need for long-term care is determined through a computerized primary judgment and a secondary judgment by a certification committee. Service applicants are then entitled to receive LTCI services according to their level of need. One of the criteria for assessing the level of need for LTCI services is the “Degree of Independence in Daily Living for the Demented Elderly,” which evaluates the level of independence considering their dementia-related symptoms and behaviors. We can extract this information from the Japanese long-term care insurance claims database, which covers LTCI users all over Japan, therefore allowing us to make nationwide comparisons among regions in Japan.

This study aimed to calculate and explore the regional difference in Dementia-free Life Expectancy (De-FLE) based on the Degree of Independence in Daily Life for the Demented Elderly. These metrics will enable us to explore whether regional disparities exist in the dementia-related independence period and what factors are associated with these trends. We expect this study to help increase De-FLE and reduce regional disparities.

## Method

### Data source

We extracted cases from the Japanese long-term care insurance claims database of the Ministry of Health, Labour and Welfare (MHLW) by the Degree of Independence in Daily Living for the Demented Elderly. This database includes information on long-term care certifications and long-term care receipts collected nationwide. We also aggregated regional variables from publicly available data sources, shown in Table 1.

**Table 1.**
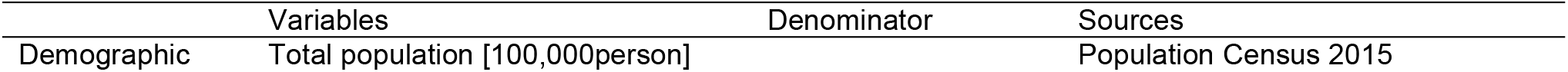
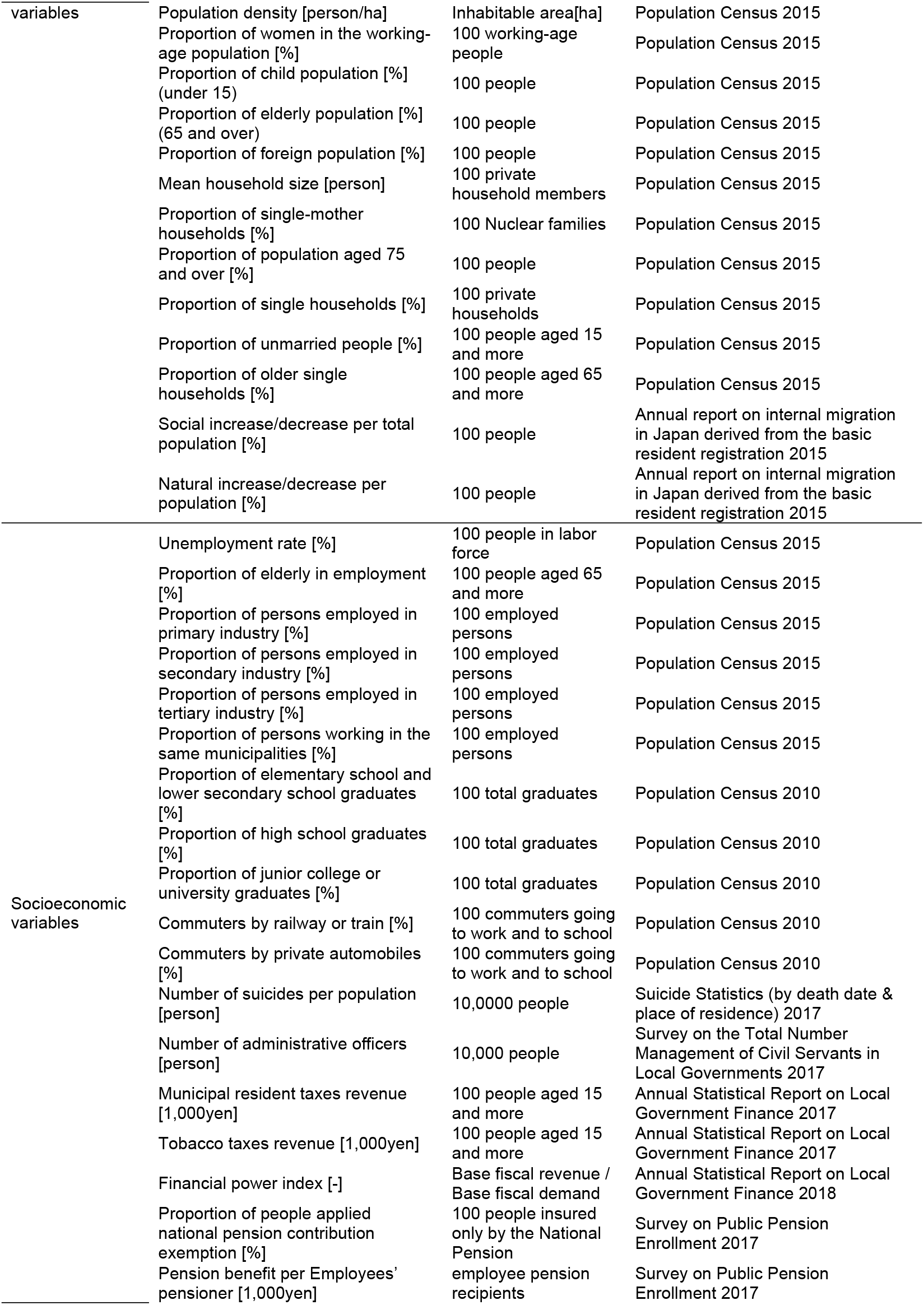
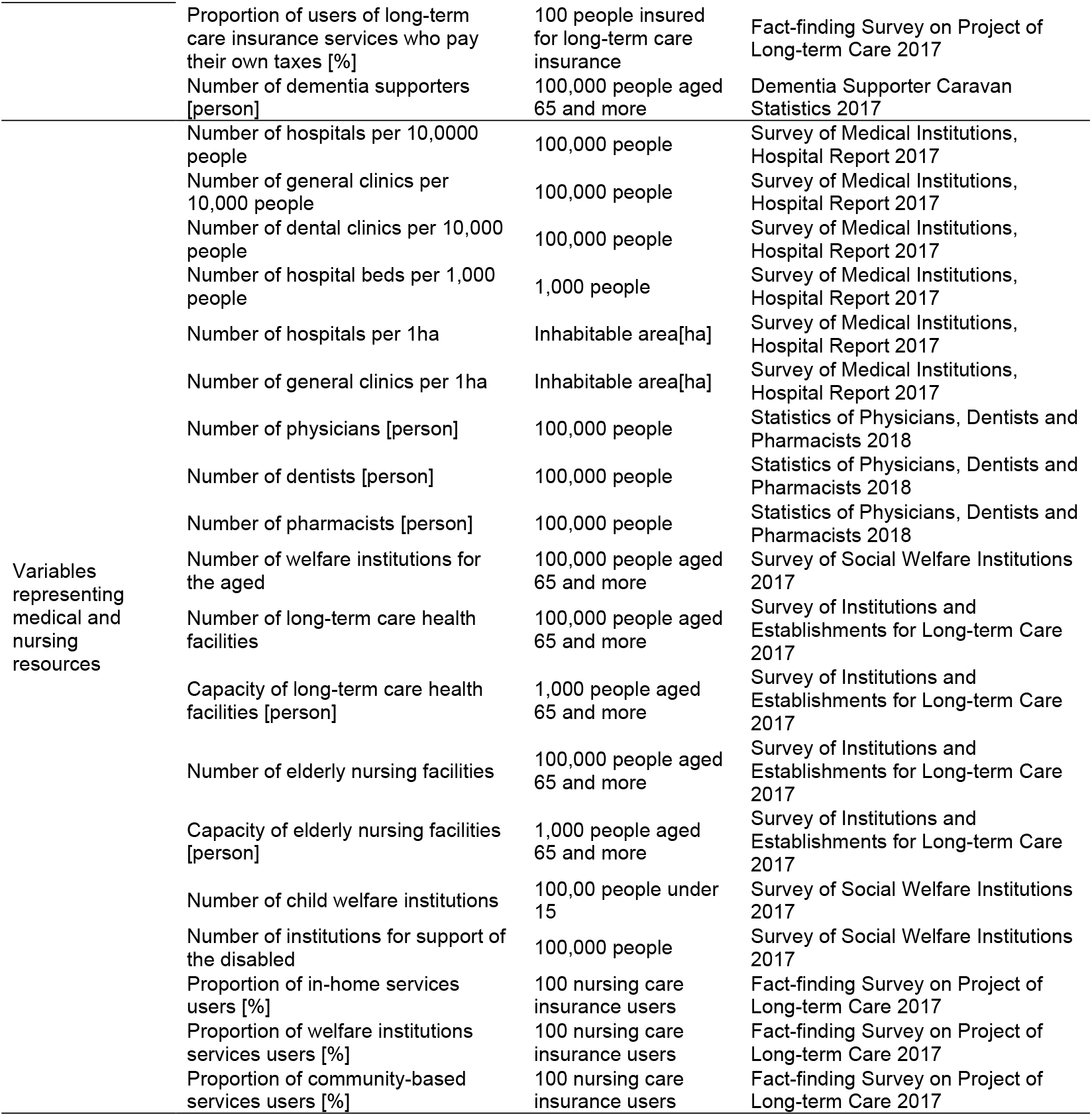
Factors related to regional characteristics, denominators used in calculations, and data sources.

### Geographical units

Secondary medical areas (SMAs), which consist of several municipalities, were used as the unit of this analysis. Japan’s Medical Care Act stipulates SMAs are the units that take comprehensive measures to foster cooperation between health care, medical care, and welfare. There are 335 SMAs in Japan, with a mean population of about 4 million in each SMA. We aggregated public statistical data by SMA and utilized it in the analysis.

### Dementia-free Life Expectancy

In our study, De-FLE refers to the period before dementia symptoms become severe enough to require support from others. We also defined LE with Dementia as the period in which people receive long-term care due to cognitive difficulties, representing the period between life expectancy and De-FLE. To calculate these metrics, we utilized the “Degree of Independence in Daily Living for the Demented Elderly,” which was created by the MHLW to facilitate objective and quick assessments by public health nurses, nurses, social workers, and care workers [13]. When the elderly in need apply for LTCI certification, investigators designated by the municipality visit their place of residence and judge their level of independence. Their family physicians also use the criteria to evaluate their independence level. The Degree of Independence in Daily Living for the Demented Elderly classifies applicants into five categories regarding their communication capacity, symptoms, and behaviors (S1. Table). In this study, we defined “unhealthy” as Independence Level II or higher to calculate De-FLE. Those assessed as Independence Level II usually live at home, but since it is challenging for them to live alone as they need support from the people around them, they often use home services in the daytime.

We extracted data from the Japanese long-term care insurance claims database in 2017 provided by the MHLW and calculated De-FLE using the number of independent degrees of daily living for the demented elderly. We defined LE with Dementia as the period between life expectancy and healthy life expectancy. The calculation followed the “Healthy Life Expectancy Calculation Program” published by the MHLW research group [14].

### Statistical analysis

We conducted a partial least squares (PLS) regression with SMA-level De-FLE and LE with Dementia for both sexes at age 65 in 2017 as the objective variables and regional factors as explanatory variables. We selected explanatory variables based on previous studies and the World Health Organization Guidelines on Dementia: “Risk Factors for Dementia,” and then aggregated them by SMAs, as shown in Table 1 [15].

PLS regression analysis is commonly used in econometrics, and in recent years, a few public health studies have adopted it [16, 17]. This model first extracts the latent components to have the largest covariance with the objective variables, then uses them to perform regression analysis. It can deal with multicollinearity among variables, making it useful for cases with many correlated explanatory variables [18]. We used leave-one-out cross-validation to identify the number of latent factors. We first identified the number of factors with a minimum predicted residual sum of squares. Then we performed Van der Vote’s test to select the smallest number of factors that had no significant difference from the above. All statistical analyses were conducted using JMP Pro 16.1 (SAS Institute Inc., Cary, NC).

This study was approved by the ethics committee of Kyoto University (approval number: R0438).

## Results

### Descriptive statistics for SMA-level LE, De-FLE, LE with Dementia, and regional variables

The mean SMA-level De-FLE at age 65 in 2017 was 17.33 years (95% confidence interval [CI] 17.27–17.38 years) for males and 20.05 years (95% CI 19.99–20.11 years) for females, a difference of 2.72 years, with standard deviations of 0.56 years and 0.54 years, respectively (Table 2).

**Table 2.**
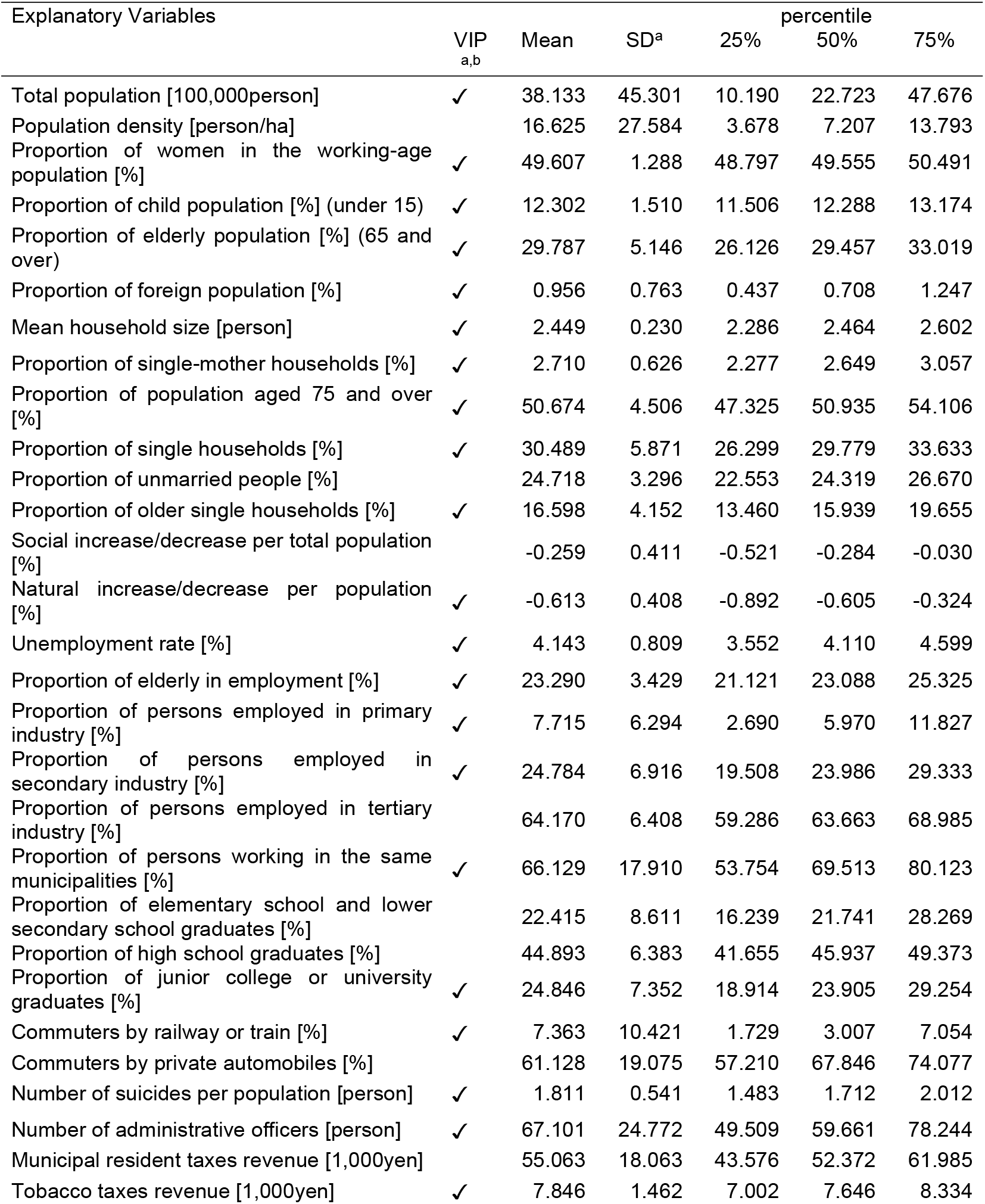
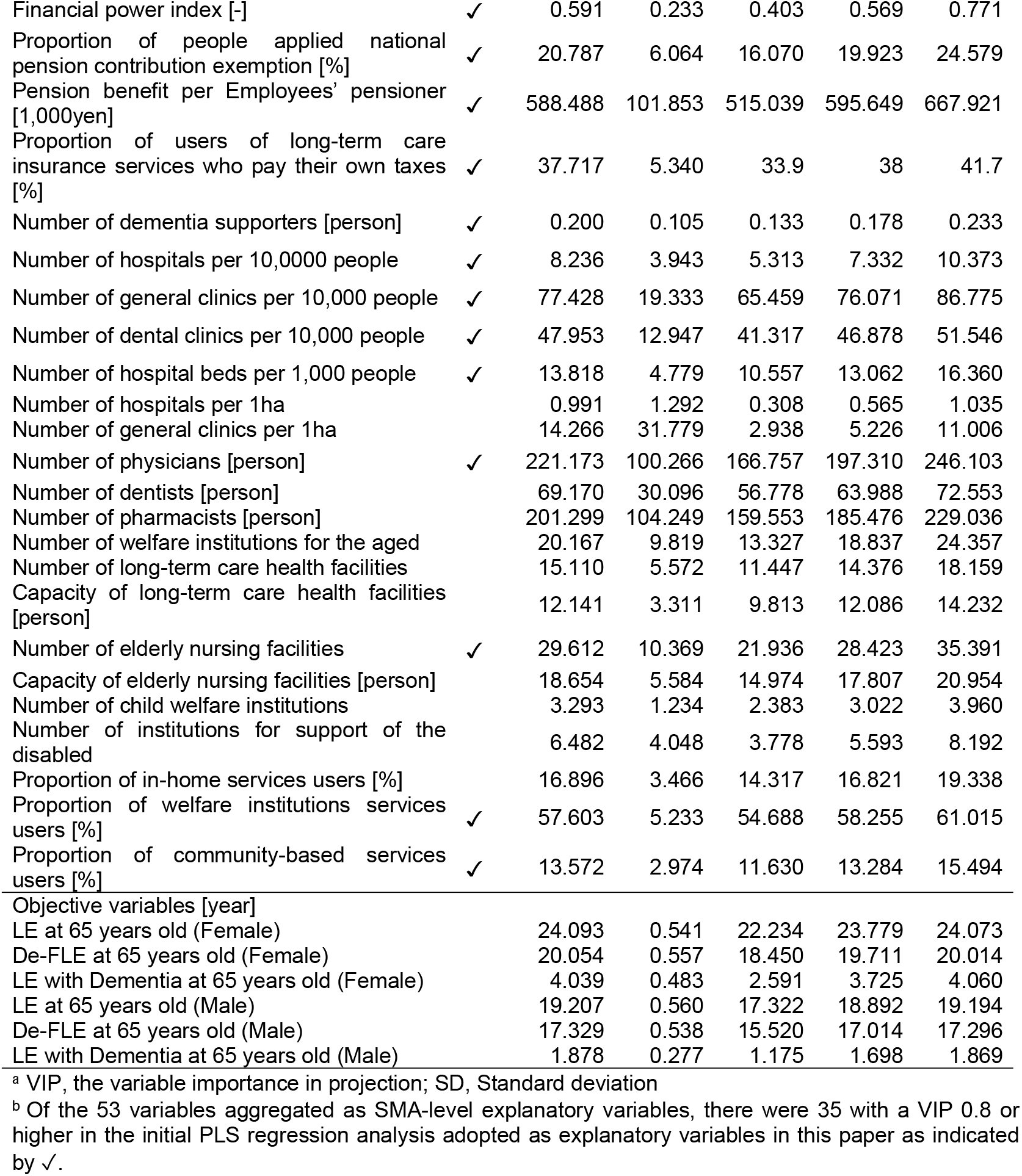
Descriptive statistics for SMA-level LE, De-FLE, LE with Dementia, and regional variables (n=333)

As for SMAs, we used data in 2017, but data submission from long-term care insurers became mandatory after 2018. In 2017, some insurers did not submit data or had significantly fewer data. To omit those insurers, we checked the list of long-term care insurers against the specific extraction data and identified 334 SMAs. We excluded one SMA with missing data from the 2015 census owing to the Great East Japan Earthquake and included 333 SMAs in the analysis.

### PLS regression analysis

Cross-validation results showed that the predicted residual sum of squares statistic was minimal when 15 factors were extracted. Then, three components were finally identified based on the outcome of Van der Vote’s Test. Factor loadings and VIPs for each of the three factors are shown in Table 3.

**Table 3.** Each component’s factor loadings/correlations and variable importance in the projection.

|  | Component<br>1 | Component<br>2 | Component<br>3 | VIP<sup>a</sup> |
| --- | --- | --- | --- | --- |
| Coefficient of determination of explanatory variables | 0.312 | 0.123 | 0.095 |  |
| Coefficient of determination of objective variables | 0.033 | 0.067 | 0.066 |  |
| De-FLE at 65 years old (Female) | -0.105 | 0.201 | 0.265 |  |
| LE with Dementia at 65 years old (Female) | 0.141 | -0.110 | 0.253 |  |
| De-FLE at 65 years old (Male) | 0.233 | 0.458 | 0.232 |  |
| LE with Dementia at 65 years old (Male) | 0.215 | -0.079 | 0.276 |  |
| Proportion of junior college or university graduates [%] | 0.924 | 0.008 | 0.045 | 0.938 |
| Natural increase/decrease per population [%] | 0.864 | -0.163 | -0.039 | 0.906 |
| Proportion of users of long-term care insurance services who pay their own taxes [%] | 0.841 | 0.271 | -0.210 | 0.807 |
| Financial power index [-] | 0.832 | -0.034 | -0.373 | 0.930 |
| Pension benefit per Employees' pensioner [1,000yen] | 0.827 | 0.120 | -0.248 | 0.824 |
| Proportion of commuters by railway or train [%] | 0.720 | -0.182 | -0.018 | 0.805 |
| Total population [100,000person] | 0.688 | -0.317 | -0.057 | 0.927 |
| Proportion of foreign population [%] | 0.617 | -0.008 | -0.183 | 0.713 |
| Number of dental clinics per 10,000 people | 0.456 | -0.397 | 0.312 | 0.942 |
| Number of physicians [person] | 0.455 | -0.285 | 0.479 | 1.002 |
| Proportion of child population [%] | 0.448 | 0.024 | 0.019 | 0.771 |
| Proportion of single households [%] | 0.304 | -0.650 | 0.386 | 1.215 |
| Number of general clinics per 10,000 people | 0.189 | -0.149 | 0.573 | 0.862 |
| Proportion of persons employed in secondary industry [%] | 0.036 | 0.504 | -0.580 | 1.180 |
| Proportion of older single households [%] | 0.027 | -0.689 | 0.448 | 1.167 |
| Proportion of women in the working-age population [%] | 0.022 | -0.206 | 0.409 | 0.903 |
| Tobacco taxes revenue [1,000yen] | -0.077 | -0.568 | 0.034 | 1.310 |
| Proportion of elderly in employment [%] | -0.098 | 0.475 | 0.177 | 0.722 |
| Number of dementia supporters [person] | -0.110 | 0.392 | 0.276 | 0.949 |
| Unemployment rate [%] | -0.122 | -0.662 | -0.248 | 1.505 |
| Proportion of community-based services users [%] | -0.147 | 0.207 | 0.381 | 0.931 |
| Mean household size [person] | -0.163 | 0.651 | -0.387 | 1.086 |
| Proportion of single-mother households [%] | -0.360 | -0.567 | 0.162 | 1.068 |
| Number of suicides per population [person] | -0.429 | -0.093 | -0.093 | 1.217 |
| Number of hospital beds per 1,000 people | -0.444 | -0.202 | 0.432 | 0.860 |
| Capacity of elderly nursing facilities [person] | -0.490 | 0.232 | 0.007 | 0.971 |
| Number of welfare institutions for the aged | -0.502 | 0.169 | 0.394 | 0.884 |
| Number of hospitals per 10,000 people | -0.574 | -0.152 | 0.417 | 1.007 |
| Proportion of in-home services users [%] | -0.631 | 0.416 | -0.108 | 0.921 |
| Proportion of people applied national pension contribution exemption [%] | -0.655 | -0.485 | 0.246 | 1.085 |
| Number of administrative officers [person] | -0.670 | 0.098 | 0.311 | 1.080 |
| Proportion of persons working in the same municipalities [%] | -0.724 | 0.010 | 0.270 | 1.022 |
| Proportion of persons employed in primary industry [%] | -0.771 | 0.119 | 0.292 | 0.839 |
| Proportion of population aged 75 and over [%] | -0.792 | 0.139 | 0.401 | 1.111 |
| Proportion of elderly population [%] | -0.835 | 0.198 | 0.162 | 1.049 |
| Proportion of elderly population [%] | -0.835 | 0.198 | 0.162 | 1.049 |

<sup>a</sup> VIP, the variable importance in projection

Component 1 was positively loaded by the proportion of junior college or university graduates, the proportion of commuters traveling by railway or train, and the financial power index. In contrast, variables such as the proportion of persons employed in primary industry and the proportion of elderly in the population loaded negatively. Thus, we named this component urbanicity. Component 2 was positively loaded by the proportion of persons employed in secondary industry and mean household size and negatively loaded by the proportion of single households, tobacco taxes revenue per population, unemployment rate, and the proportion of single-mother households. We defined this as the socioeconomic conditions-related component. Component 3 was characterized by positive loadings on the number of general clinics per 100,000 people and the number of hospital beds per 1,000 people, and thereby named the healthcare resources component.

The correlation and loading plot of variables for components 1 and 2 is shown in Fig 1(A). De-FLE for males and LE with Dementia were positively correlated with the score of component 1. De-FLE for both sexes was positively correlated with the score of component 2, especially for males. Of the three components, component 1 had the widest variation of explanatory variables, but its variation in objective variables was smaller than that of component 2. On the other hand, tobacco taxes revenue per population and unemployment rate had high negative loadings on component 2 but nearly zero loading on component 1. The correlation and loading plots of components 2/3 and 1/3 are shown in Fig 1(B) and 1(C), respectively. With component 3, all objective variables were positively correlated. The variation in objective variables of components 1 and 3 was much smaller than that of component 2.

**Fig 1.**
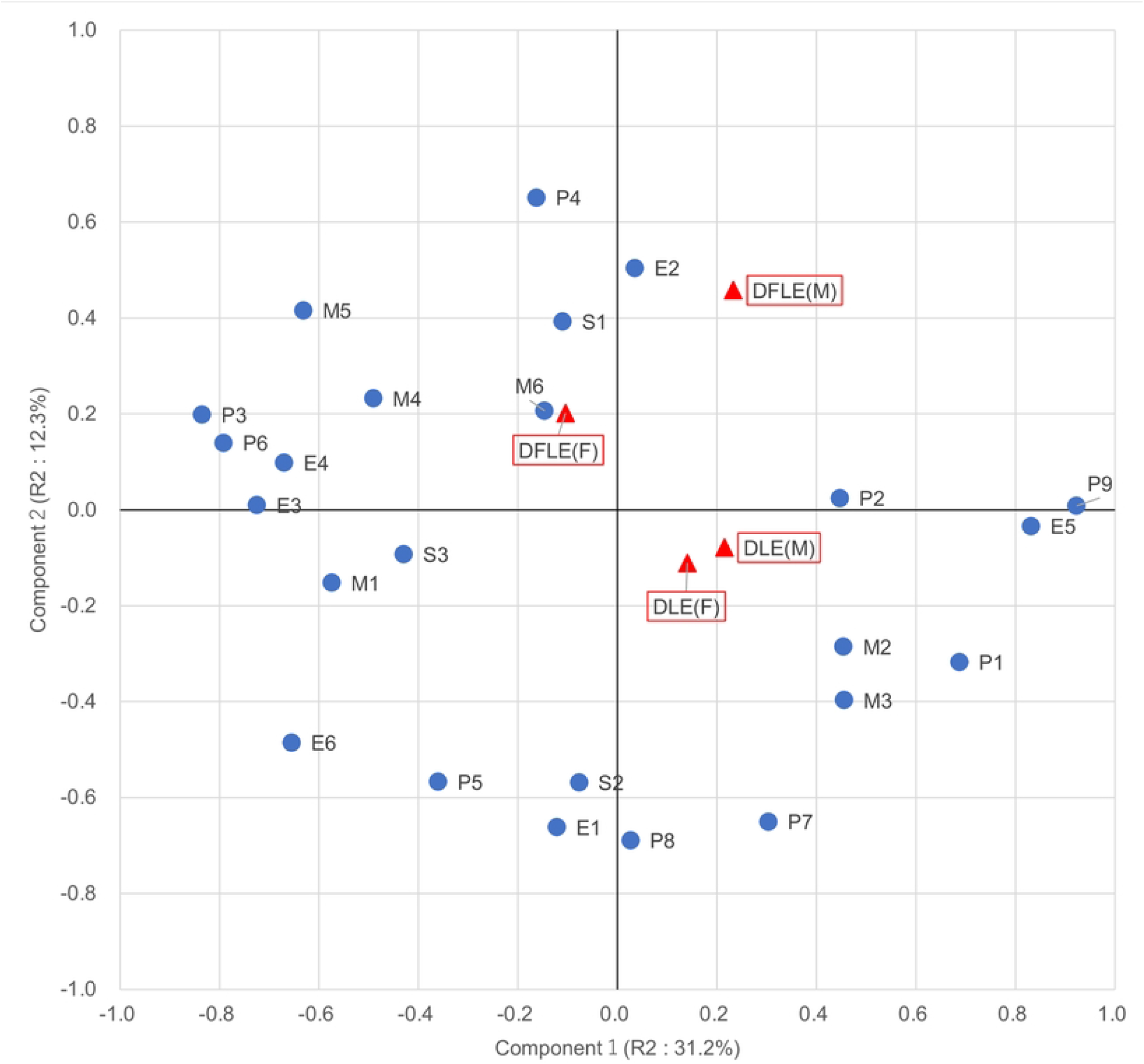
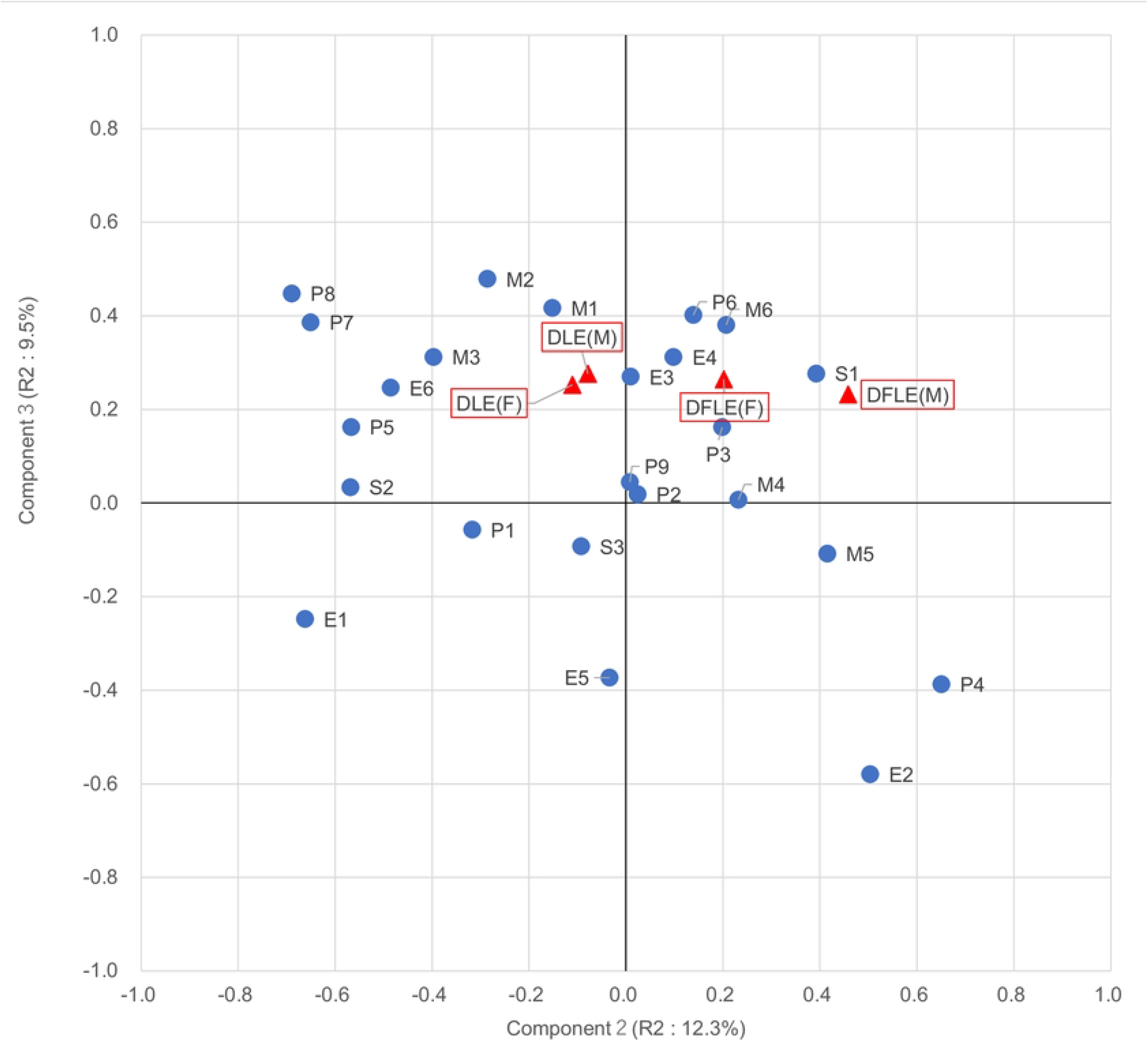
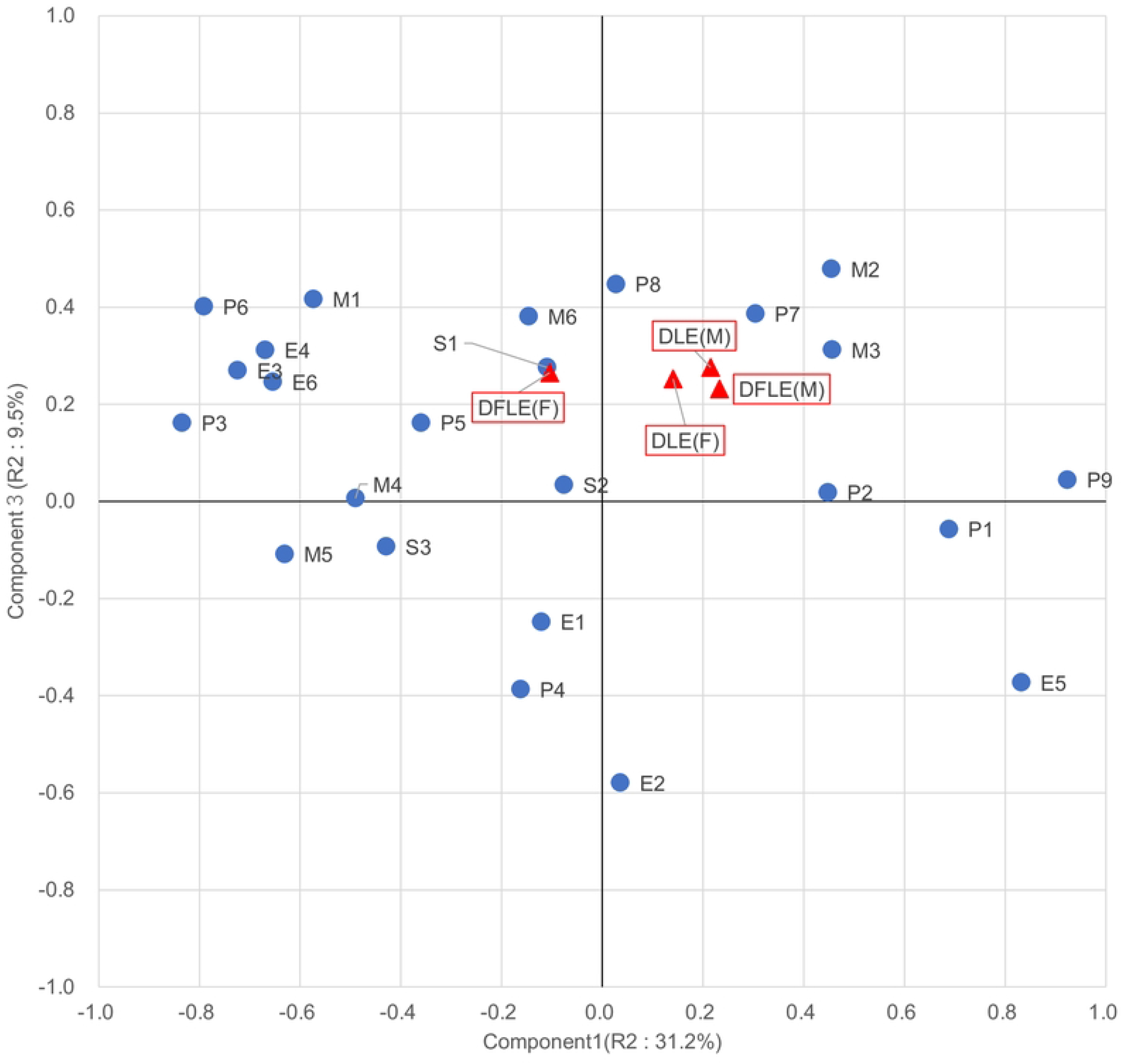
Correlation and loading plots for the first three components. (A) Component 1 and component 2; (B) Component 2 and component 3; (C) Component 1 and component 3 Red triangles indicate the loadings of objective variables, and blue circles indicate the loadings of the explanatory variables. P1, Total population; P2, Proportion of child population; P3, Proportion of elderly population; P4, Mean household size; P5, Proportion of single-mother households; P6, Proportion of population aged 75 and over; P7, Proportion of single households; P8, Proportion of older single households; P9, Proportion of junior college or university graduates; S1, Number of dementia supporters per elderly population; S2, Tobacco taxes revenue per population; S3, Number of Suicides per population; E1, Unemployment rate; E2, Proportion of persons employed in secondary industry; E3, Proportion of persons working in the same municipalities; E4, Number of administrative officers; E5, Financial power index; E6, Proportion of people applied national pension contribution exemption; M1, Number of hospitals per 100,000 people; M2, Number of physicians; M3, Number of dental clinics per 100,000 people; M4, Capacity of elderly nursing facilities; M5, Proportion of welfare institutions services users; M6, Proportion of community-based services users; DFLE(F), De-FLE (Female); DLE (F), LE with Dementia (Female); DFLE(M), De-FLE (Male); DLE(M), LE with Dementia (Male)

## Discussion

In this study, we calculated SMA-level De-FLE and LE with Dementia for both sexes and then conducted PLS regression analysis to examine regional disparities in Japan. De-FLE and LE with Dementia varied among SMAs, and mean SMA-level De-FLE was longer for women than men. Three latent components were identified, which represented urbanicity, socioeconomic conditions, and healthcare resources. Socioeconomic conditions explained De-FLE the best.

Component 1 can be interpreted to represent urbanicity since it was associated with variables such as a high population and a low proportion of elderly in the population, which are typical of urban areas. The high score of component 2 represents good socioeconomic conditions as it was characterized by negative loadings for unemployment rate, the proportion of unmarried people, and tobacco taxes revenue per population. The loadings of these variables on component 1 were nearly zero. The high score on component 3 represents rich healthcare resources because it had positive loadings for the number of general clinics per population and the number of hospital beds per 1,000 people.

While component 1 explains the region’s characteristics well, its contribution to De-FLE can be described as small, given that the coefficient of determination for the explanatory variable was as large as 31.2%, contrary to the objective variable, which was as small as 3.28%. Component 1 was positively correlated with De-FLE for males, but it was not correlated with De-FLE for females. Previous research also showed that rural centenarians enjoyed better health status than their urban counterparts [19]. The score of component 2 had a positive correlation to De-FLE. Therefore, De-FLE will be longer when living in better socioeconomic conditions. Component 2 can be referred to as the most crucial component in explaining LE with Dementia because it had the largest coefficient of determination for the objective variables of the three, thus explaining De-FLE. This result was consistent with the well-known fact that maintaining good socioeconomic status is essential to staying healthy [20]. Improving living conditions will prolong De-FLE. The score of component 3 was positively correlated with De-FLE, suggesting that access to healthcare resources was also positively associated with these metrics. The development of a healthcare delivery system may result in longer De-FLE [21, 22].

The score of component 1 was positively correlated with LE with Dementia for both sexes. Several studies in the US have reported higher mortality among people living in non-metropolitan than in metropolitan areas [23, 24]. Component 2 had a negative correlation with LE with Dementia, contrary to De-FLE. Socioeconomic conditions may affect De-FLE more than LE with Dementia.

The score of component 3 was positively correlated with both De-FLE and LE with Dementia, suggesting that access to healthcare resources was also positively associated with these metrics. Component 3 was positively correlated with LE with Dementia. A large number of people with dementia might bring about the need to develop a healthcare delivery system, thereby leading to an increase in physicians. However, a causal relationship remains unclear, and further research is warranted.

Component 2 was positively correlated with De-FLE and contributed to a higher degree than components 1 and 3, as shown in Table 3. This means socioeconomic conditions best explain the variation in De-FLE, compared with urbanicity and healthcare resources. Most of the variables that illustrate component 2 well had little loadings on component 1, as shown in Fig 1. For example, variables with high negative loadings on component 2 were the unemployment rate, the proportion of single households, tobacco taxes revenue per population, mean household size, and the number of dementia supporters per elderly person in the population. These variables had little loadings on component 1, meaning that these variables were positively associated with De-FLE regardless of urbanicity. On the other hand, variables with high negative loadings on component 2 were the mean household size and the number of dementia supporters per elderly person in the population, implying that they were also positively associated with De-FLE. These also had little loadings on component 1, which means that they are negatively associated with De-FLE in both urban and rural areas. We will further discuss socioeconomic conditions below.

The unemployment rate was a critical variable of component 2. A previous study suggested a significant relationship between work and self-rated health among older adults [25]. A sense of fulfillment is also necessary because working only for financial gain diminishes the health benefits of working after retirement [26]. Measures to reduce the unemployment rate by sufficiently arranging rewarding jobs might work toward extending De-FLE.

Tobacco taxes revenue per population was also a crucial variable of socioeconomic conditions. The adverse effects of tobacco on overall health, not just dementia, have led to the enforcement of many tobacco control and policy measures [27, 28]. Social factors such as poor housing conditions, low wages, isolated parents, unemployment, and homelessness are closely linked to high rates of smoking and low rates of smoking cessation, and smoking is a significant cause of illness and premature death [29]. Given the above report and our study, smoking will not only directly but indirectly harm a person’s health status and De-FLE.

The number of dementia supporters per elderly person in the population also explains the characteristics of socioeconomic conditions. Previous studies revealed that extensive community resources contribute to longer healthy life expectancy [30, 31]. Improved initiatives, such as introducing dementia supporters, may play a critical role in encouraging government and community residents to better understand people with dementia, which will help prevent isolation among the elderly [32].

Variables representing family structure, such as the proportion of older single households and mean household size, were influential factors in explaining socioeconomic conditions. Previous research has found that household size was negatively associated with mortality from dementia [33]. Another previous study noted that people who live alone are more likely to move into a nursing home than those who live with a family member [34]. Our study suggests that household size may also be negatively associated with the number of people needing care due to dementia. However, it is estimated that the proportion of single households will continue to increase, accounting for 40% of all households by 2040 [35]. Given this social trend, promoting the development of community-based support systems will be imperative to avoid burdening family members and ensure that everyone receives appropriate care.

As for gender differences, the correlation between component 1 and De-FLE was positive for males and nearly zero for females, which means urbanicity could affect males and females differently. Previous studies on the association between city size and health showed that areas with higher urbanicity had more people aged 100 years and over and a lower risk of physical disability [36-38]. In addition, Component 2, which represents socioeconomic conditions, correlated with De-FLE better for males than females. Several reports have found gender differences in the association between socioeconomic conditions and healthy life expectancy [39, 40]. This study found that socioeconomic conditions explain De-FLE particularly well for males, suggesting the need for community support considering gender.

## Strengths and limitations

We examined regional disparities in the time before the elderly needed substantial long-term care. Using LTCI data and official statistical data allowed us to make comparative observations not only for specific regions but also across all regions of Japan. On the other hand, there are some limitations to this study. First, an ecological fallacy, a phenomenon in which observed associations true at the population level do not apply at the individual level, might occur in this study. We considered variables reported to associate with individual-level health status in previous studies and found consistent results. In addition, this study aimed to identify factors associated with regional differences in De-FLE rather than to explore individual-level factors. Furthermore, while region-level statistics may not apply to the population over 65 years of age, we consider the region-level variables in this study as a neighborhood environment rather than a nature that directly describes that population. Finally, some official statistical data, such as educational background and commuting patterns, were collected from the 2010 Census results, and the situation may have changed since that time. Continuous monitoring with new data is warranted.

## Conclusions

This study calculated Dementia-free Life Expectancy at the secondary medical area level as a new indicator for understanding the regional statuses of the elderly with dementia across the entire country of Japan and explored its association with regional variables. We found that socioeconomic conditions correlate with Dementia-free Life Expectancy more than urbanicity and healthcare resources factors.

## Data Availability

Data from the Japanese long-term care insurance claims database cannot be shared publicly because the authors do not have permission. Data we used in aggregating regional variables are available on “e-Stat,” a portal site for Japanese Government Statistics. https://www.e-stat.go.jp/en

## Supporting information

S1 Table. Degree of Independence in Daily Living for the Demented Elderly

